# Machine learning-based neuroimaging for prediction of deep brain stimulation outcomes in movement disorders: Systematic review and meta-analysis

**DOI:** 10.64898/2026.07.22.26358674

**Authors:** Mohammad-Jasim Golzarian, Shahryar Rajai Firouzabadi, Zanyar Hajiesmaipoor, Zahra Azizan, Alireza Alikhani, Pouria Moshayedi

**Author notes:** **Corresponding author:** Mohammad-Jasim Golzarian, MD, School of Medicine, Shahid Beheshti University of Medical Sciences, Tehran, Iran.

## Abstract

**Background:** Deep Brain Stimulation (DBS) surgery is a treatment of choice for movement disorders, and utilizes an implanted electrical pulse generator that administers electrical stimulation to designated brain regions responsible for motor control. The preoperative identification of effective predictive factors is of utmost importance for appropriate patient selection. In this study, we evaluate the potential of machine learning-based neuroimaging for predicting DBS outcomes. (PROSPERO Registration: CRD420261279318)

**Method:** Following the PRISMA statement, eligible studies were selected through searching three databases (PubMed, Scopus, Web of Science) on November 6, 2025. Methodological quality was assessed using the PROBAST+AI tool. Random-effects models pooled discrimination performance (AUC). Heterogeneity was investigated using meta-regressions for age and gender alongside subgroup analysis by type of algorithm. Publication bias was assessed using Egger’s regression test.

**Results:** Twenty studies were included in the analysis. Most investigations focused on PD, STN-DBS, and postoperative motor improvement, while a smaller number assessed neuropsychiatric outcomes. Overall, the pooled discrimination for models predicting motor outcomes showed an AUC of 0.86, and the pooled models for delirium showed an AUC of 0.87. Regarding the risk of bias assessment, seven studies were classified as low risk, while thirteen were identified as high risk.

**Conclusion:** Machine learning-based neuroimaging shows promising potential for preoperative prediction of DBS outcomes. However, the current literature is characterized by a persistent gap between encouraging discrimination and reliable clinical readiness. The main weakness of the field lies in analytical rigor and generalizability. These models should currently only be considered as promising research tools.

## 1. Introduction

Machine learning (ML) constitutes a computational paradigm for recognizing patterns in datasets ^1^. It has exerted a profound effect on clinical environments, ranging from clinical decision support systems to surgical assistance ^1,2^. One notable application of this technology could be in Deep Brain Stimulation (DBS) surgery, which represents a highly complex surgical procedure and treatment of choice for movement disorders, such as Parkinson’s disease (PD), dystonia, and essential tremor ^2,3^. DBS utilizes an implanted electrical pulse generator that administers electrical stimulation to designated regions of the brain, which are responsible for motor control ^2^. The successful implantation and subsequent programming of the electrodes are notably complex tasks that necessitate interdisciplinary collaboration among neurology and neurosurgery specialties ^2,3^. Notably, despite the invasiveness of the surgical procedure, many DBS patients do not exhibit symptomatic improvement ^2,3^. In certain cases, DBS may even worsen symptoms ^2,4^. Occasionally, further invasive procedures become necessary to either remove or reposition the pulse generator ^2,4^. The efficacy of DBS relies on several factors, including preoperative patient candidacy, accurate targeting during the implantation phase, meticulous postoperative programming, and continuous medical management ^5,6^. While precise targeting during implantation and optimal postoperative programming are crucial for achieving favorable outcomes, considerable individual variability exists in the postoperative amelioration of symptoms ^6^. Hence, the preoperative identification of effective predictive factors is of utmost importance for appropriate patient selection ^6^. The levodopa challenge test (LCT) is the most widely used predictive factor for assessing motor prognosis in DBS ^6,7^. However, its predictive accuracy regarding long-term postoperative motor outcomes exhibits significant variability and a lack of precision ^6,7^. Furthermore, the LCT has additional drawbacks, including increased risk of motor complications from levodopa cessation for at least 12 hours, exacerbation of drug-induced dyskinesia with levodopa use, and the time-intensiveness of this challenge ^8^. Additionally, the LCT is inadequate in predicting levodopa-resistant symptoms, such as improvements in tremor and specific dyskinesias ^9^. Further inaccuracies in LCT results stem from its dependence on dosage and the duration of the disease, which may induce challenging withdrawal symptoms ^9^. Numerous investigations have endeavored to find reliable supplementary predictors of DBS responsiveness, with varied success ^10^. Age has been identified as a negative prognostic indicator for subthalamic nucleus (STN) DBS; however, more recent studies have indicated that there is no significant difference in DBS responsiveness or complication rates solely based on age ^10–12^. Similarly, while tremor has been documented to respond particularly consistently to STN DBS, the subtypes of PD characterized by postural instability-gait difficulty and tremor dominance do not appear to exhibit significant differences in their overall motor responses ^10^. Limitations in the predictive capacity of clinical characteristics regarding DBS responsiveness have prompted investigations utilizing brain structural and functional neuroimaging modalities to correlate DBS efficacy ^10^. Various volumetric MRI studies have established associations between structural cerebral differences and DBS outcomes through the examination of both cortical and subcortical volumetric metrics, including ventricular, thalamic ^13^, and midbrain volumes ^14^, and visuo-motor ^15^, superior frontal, and paracentral cortical thickness ^10,16^.

Moreover, studies have been conducted to assess the relationship between neuronal activity (potentially influenced by PD), which is altered by DBS, and the resultant outcomes of DBS ^17,18^. Nevertheless, the interpretation and use of features extracted from medical imaging could be challenging and expertise-dependent ^19^. Recent improvements in ML and medical imaging have shown promise in solving these problems ^20^. Besides, the trainability of ML models on existing data and making outcome predictions for unseen cases as its strength over traditional statistics makes it increasingly used in clinical practice ^19^.

In this systematic review and meta-analysis, for the first time, we evaluate the role of ML-based neuroimaging in outcome prediction for DBS.

## 2. Methods and Materials

### 2.1. Search Strategy and Eligibility Criteria

Search strategy: A comprehensive literature search was conducted using three online databases, PubMed, Scopus, and Web of Science on November 6, 2025. The following search keywords were used: “Deep Brain Stimulation” or “deep brain stimulation” or “DBS” and “Movement Disorders” or “Parkinson Disease” or “Essential Tremor” or “Dystonia” or “Tremor” or “movement disorder*” or “parkinson*” or “essential tremor” or “dystoni*” or “tremor*” and “Machine Learning” or “Artificial Intelligence” or “machine learning” or “deep learning” or “neural network*” or “artificial intelligence” or “AI” or “random forest” or “support vector machine*” or “SVM” or “logistic regression” and “Magnetic Resonance Imaging” or “Diffusion Tensor Imaging” or “Positron-Emission Tomography” or “MRI” or “fMRI” or “DTI” or “diffusion” or “PET” or “brain imaging” or “neuroimaging” and “predict*” or “outcome” or “candidat*” or “response” or “prognosis”.

We included all original English language studies conducted on patients who underwent DBS that used ML/AI algorithms for outcome prediction utilizing neuroimaging data and reporting outcomes with quantitative measures. We excluded studies with non-human subjects, in vitro experiments, case reports, and patients with movement disorders who did not undergo deep-brain stimulation. Review studies, editorials, conference abstracts without full-text, non-English publications, studies focusing only on targeting without outcome prediction, and studies without neuroimaging data or without the use of machine learning-based predictive models will also be excluded.

### 2.2. Study Selection

Two reviewers (Sh.R. and Z.H.) independently screened the titles and abstracts of all identified records for eligibility. Studies deemed potentially relevant were subsequently assessed through full-text review. Disagreements between reviewers were resolved by discussion, and if necessary, by consultation with a third reviewer (M.G.).

### 2.3. Data Extraction

Two reviewers (A.A. and SH.R.) independently extracted demographic, clinical, and ML-related data of each included study through full-text review. These extracted data were checked by an independent reviewer (M.G.) for possible mistakes. For missing data, supplementary files were explored. (See Table 1 and Supplemental Table 1.)

### 2.4. Quality assessment

The risk of bias and applicability of included prediction model studies (development, validation, or update) were independently assessed by two reviewers (Z.A. and Z.H.) using the Prediction model Risk Of Bias ASsessment Tool for Artificial Intelligence (PROBAST+AI), the updated version of PROBAST specifically developed for prediction models using regression or artificial intelligence/machine learning methods ^21^. PROBAST+AI evaluates four domains (Participants and data sources, Predictors, Outcome, and Analysis) with targeted signaling questions, leading to domain-level and overall judgments of low, high, or unclear risk of bias, as well as concerns regarding applicability to our review question by focusing on neuroimaging-based ML models for predicting DBS outcomes in neurological disorders.

Disagreements were resolved by discussion or consultation with a third reviewer (M.G.). Results are summarized in graphical form (e.g., traffic light plots and summary bar plots) for each domain and overall.

### 2.5. Data synthesis

We performed a meta-analysis of model performance metrics where data were sufficiently homogeneous and amenable to pooling, however, due to substantial clinical, methodological, and statistical heterogeneity across some of the included studies—including differences in machine learning algorithms (e.g., support vector machine (SVM), logistic regression (LR), random forest (RF)), neuroimaging modalities (e.g., fMRI, DTI, structural MRI), predictor features, outcome definitions (e.g., motor improvement, response rate, Unified Parkinson’s disease rating scale (UPDRS) change), validation strategies (internal vs. external), sample sizes, and reporting of performance metrics (e.g., area under the curve (AUC), accuracy, sensitivity/specificity, calibration)—a meta-analysis including all studies was deemed inappropriate. Pooled estimates, forest plots, and measures of heterogeneity (e.g., I² and τ²) are presented for meta-analyses (see Results). For the remaining studies (and to contextualize the meta-analysis results), we conducted a structured narrative synthesis of all quantitative findings to avoid relying solely on pooled estimates. Studies were grouped based on types of outcomes, according to predefined criteria relevant to the review question. We highlighted patterns of agreement or variation, and explored potential sources of heterogeneity (e.g., risk of bias per PROBAST+AI, external validation status, sample characteristics). Visual aids such as summary tables of performance metrics across all studies were used to facilitate comparison. Limitations of the partial meta-analysis and narrative synthesis (e.g., potential subjectivity in grouping, reduced precision for non-pooled studies) are discussed in the discussion section.

### 2.6. Statistical Analyses

All statistical analyses were conducted by Stata 18. For the study of Yang et al. ^22^, where the SE for AUC was not directly reported, it was estimated using the nonparametric method proposed by Hanley & McNeil ^23^, which is suitable for continuously distributed data in neuroimaging-based prediction models. We only used values from test groups, not training groups. Performance was evaluated using reported AUC, R2, or MSE. Meta-analysis was based on AUC (with logit transformation) as it was the most consistently reported discrimination metric. Other metrics (R^2^, MSE) were narratively synthesized due to heterogeneity in reporting. Precomputed AUC and its respective standard error (SE) were the only acceptable data entry forms, and 95% confidence intervals were converted to SE. A random effects model was employed due to differing validation methods and algorithms. Heterogeneity was assessed using the I2 statistic, with an I21>150% signifying substantial heterogeneity. A p-value of <10.05 was considered statistically significant. Heterogeneity was investigated using meta-regressions for age and gender (male ratio) alongside subgroup analysis by type of algorithm. Other covariates, such as the type of features, were either too homogeneous or inconsistently reported to allow for meta-regression or subgroup analysis. Publication bias was assessed using Egger’s regression test.

## 3. Results

Our online database search resulted in 402 articles, of which 257 were selected for title and abstract screening after the removal of 145 duplicates. 42 were chosen for full-text evaluation, and 20 studies were included in this review. Reasons for exclusion in the second phase of screening include not reporting outcome, focusing only on targeting, without neuroimaging data, without ML use, and not discriminating with or without neuroimaging data (Figure 1). The overall study selection process followed PRISMA guidelines and is illustrated in the PRISMA flow diagram ^24^ (Figure 1).

**Figure 1:**
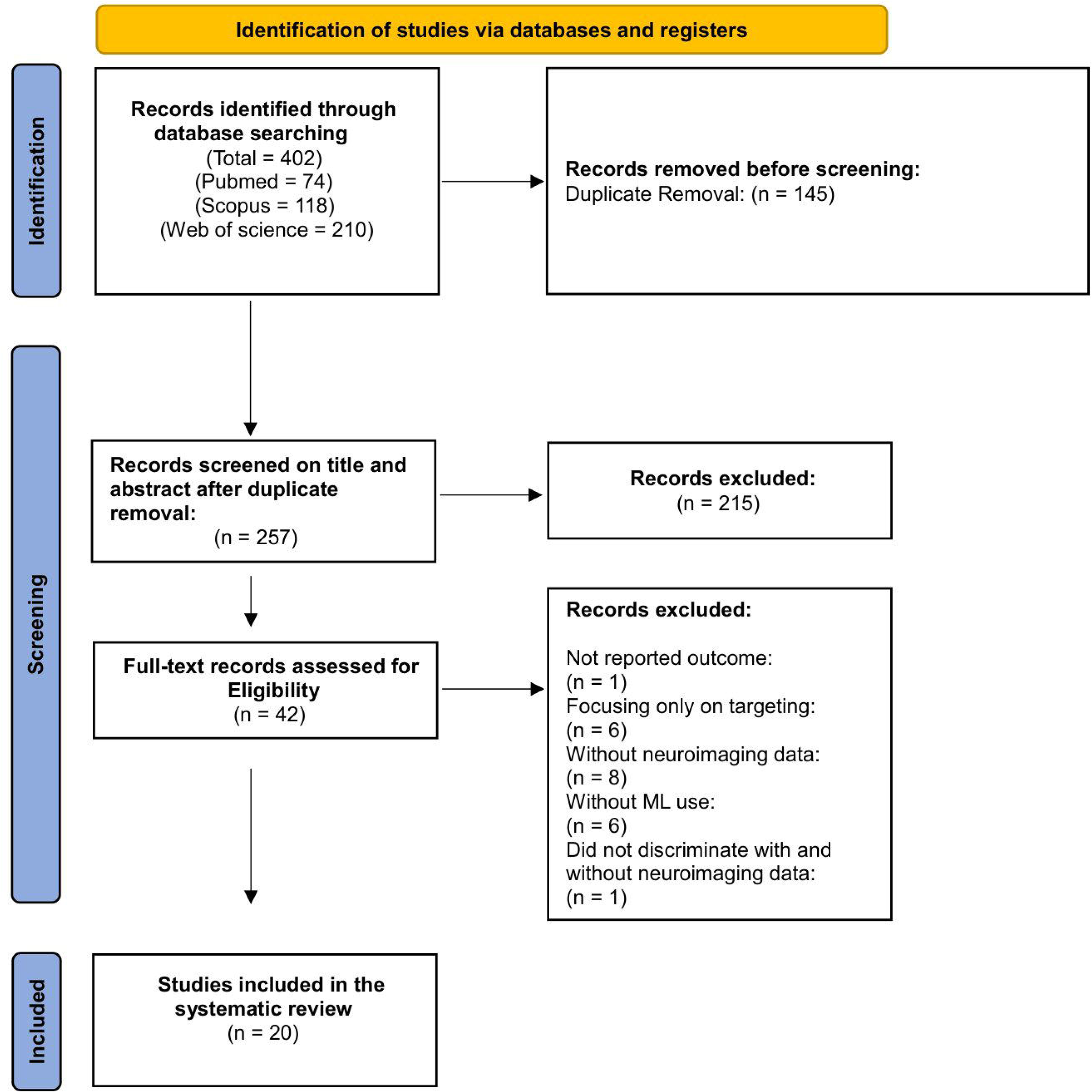
PRISMA flowchart.

### 3.1. Study Characteristics

The 20 included studies were published between 2019 and 2025, with eleven studies (55%) being published since 2023. Most studies were conducted in China (n=10), followed by the USA (n=3) and Lithuania (n=2), with one study each in France, Germany, India, South Korea, and the UK. The cause for DBS was PD in all studies, except one, which included patients with dystonia. ^25^ The number of patients undergoing DBS varied between 33 ^8,26^ and 209 ^27^, and a total of 1,551 patients were included in this review. The average age of participants ranged from 51.3 ^26^ to 64 ^27^, with male ratios between 44.7% ^28^ to 67.5%. ^19^ DBS target was mostly STN (n=16), followed by Globus Pallidus internus (Gpi) (n=4), and Ventral Intermediate nucleus (VIM) (n=1). Postoperative outcomes investigated were mostly motor function assessed via UPDRS III (n=17) with dystonia rating, delirium, freezing of gait, Hoehn & Yahr (H-Y) stage, UPDRS I, UPDRS II, depression, and cognitive function, each being investigated by one study.

The neuroimaging modalities analyzed were mostly MRI (n=14), followed by fMRI (n=6), DTI (n=2), and quantitative susceptibility mapping (QSM) (n=1). Further characteristics are available in Table 1.

### 3.2. AI Model Characteristics

53 models were developed across 20 studies. A wide range of algorithms were employed, with the most popular being SVM (n=14) and logistic regression (LR) (n=7). Features used to train the models were mostly radiomics, followed by functional connectivity. The most common validation methods were K-fold cross-validation and train-test without bootstrapping. Performance of the models varied significantly, with sensitivities ranging from 0% ^29^ to 100% ^29^, specificities ranging from 33% ^29^ to 100% ^29^, and AUCs ranging from 30% ^9,30^ to 99% ^9,29^. The best performing models were mostly based on an LR (n=5) or SVM (n=5) algorithm. Further details regarding model performance and characteristics are available in Supplemental Table 1.

### 3.3. Performance of AI models in Predicting Postoperative Outcomes

#### 3.3.1. Motor Function

Among studies predicting postoperative motor function, six studies ^19,20,22,30–32^ consisting of twenty models had sufficient information for meta-analysis. By pooling the twenty models, we found an AUC of 86% (95% CI: 79%, 90%) with high heterogeneity (I^2^: 99.8%) and significant publication bias (Egger’s p-value 0.002). Subgroup analysis by type of algorithm found significant between-group differences (p-value<0.01). Meta-regressions by age (coefficient = −0.021, p-value = 0.873, R^2^ = 0%) and male ratio (coefficient = 2.94, p-value = 0.195, R^2^ = 3.6%) were insignificant. Features utilized by the models were mostly radiomics, with the exception of Yang et al. ^22^, which used amplitudes of low-frequency fluctuations and Chang et al. ^20^, which combined radiomics with multi-instance learning (Figure 2).

**Figure 2:**
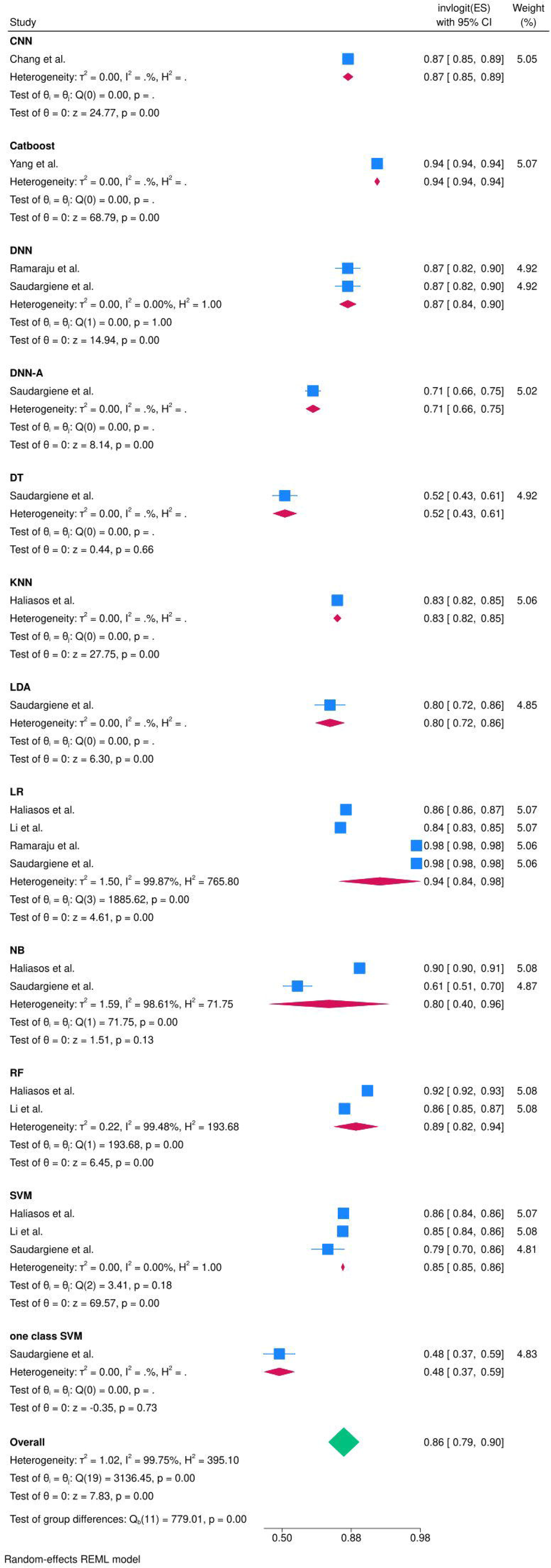
Meta-analysis of AUC of models predicting postoperative motor function.

Among studies not included in the meta-analysis, Chang et al. ^28^ developed a SVR model utilizing serum uric acid related brain functional connectivity with a mean standard error (MSE) of 0.173, Chen et al. ^33^ made an SVM model using brain morphology parameters with an R^2^ of 0.3224, Chen et al. ^34^ developed an SVM model using structural and functional connectivity with an R^2^ of 0.2478, Diao et al. ^6^ developed an XGboost model using a structural covariance network with an AUC of 80% and an R^2^ of 0.67. Liu et al. ^8^ developed two models using LR, one utilizing radiomics and the other combining radiomics with a levodopa challenge test with AUCs of 85% and 83%, respectively. Peralta et al. ^35^ developed four SVM models using preoperative clinical scores selected by an artificial neural network (ANN) to predict postoperative motor function both during the OFF and ON stage, with correlation coefficients ranging from 0.3 to 0.5 and 0.3 to 0.4, respectively. The models differed by the postoperative time point of assessment, with poorer performance in predicting OFF stage motor function 3 years after DBS compared to 3 months afterwards and better performance in predicting ON stage motor function 1 year after DBS compared to 3 months afterwards. Mo et al. ^26^ developed an SVM model using surface-based morphometry with an R^2^ of 0.14. Roberts et al. ^9^ developed a Least Absolute Shrinkage and Selection Operator (LASSO) regression based on radiomics with an AUC of 99% once a noise-compensated dataset was used for training the model. Shang et al. ^36^ developed five models utilizing functional connectivity, and the MSE of the models ranged from 240 for a gradient boosting machine model to 862 for a linear regression model. Wang et al. ^37^ developed four ridge regression models utilizing functional connectivity based on inter- or intra-hemispheric asymmetry and selected by an RF model. The model using intra-left hemispheric functional connectivity has the best performance with a correlation coefficient of 0.37, while the inter-hemispheric heterotopic-based model had the lowest correlation coefficient of 0.08.

Lastly, Younce et al. ^10^ developed a LASSO regression model using a combination of functional connectivity, brain morphology, and clinical variables with an R^2^ of 0.3153. Overall, the performance of the models varied significantly by the type of algorithm used, yet most had AUCs of 80% and R^2^ of at least 0.30.

Four studies combined their neuroimaging features with clinical or demographic variables ^8,10,28,30^. The resulting models generally had acceptable performance with AUCs of 66% ^30^ to 87% ^28^, and an R^2^ of 0.31^10^. Two studies also compared models combining neuroimaging data with those based exclusively on neuroimaging, and found models solely using neuroimaging performed slightly better or were similar in comparison ^8,30^.

#### 3.3.2. Neuropsychiatric Outcomes

Two studies developed models to predict neuropsychiatric outcomes, with one study developing multiple models to predict postoperative delirium ^29^ and another developing SVM models to predict apathy, anxiety, depression, and cognitive function after DBS ^35^. The models predicting delirium were pooled to show an AUC of 87% (95% CI: 71%, 95%) with high heterogeneity (I^2^: 99.9%). Subgroup analysis found a significant difference between models by the employed algorithms (p-value<0.01). Publication bias and meta-regressions by age and gender were not feasible due to the inclusion of only one study (Figure 3). Features utilized were exclusively radiomics. The models predicting apathy, anxiety, depression, and cognitive function could not be meta-analyzed but had mean correlation coefficients of 0.4, 0.3, 0.3, and 0.4, respectively.

**Figure 3:**
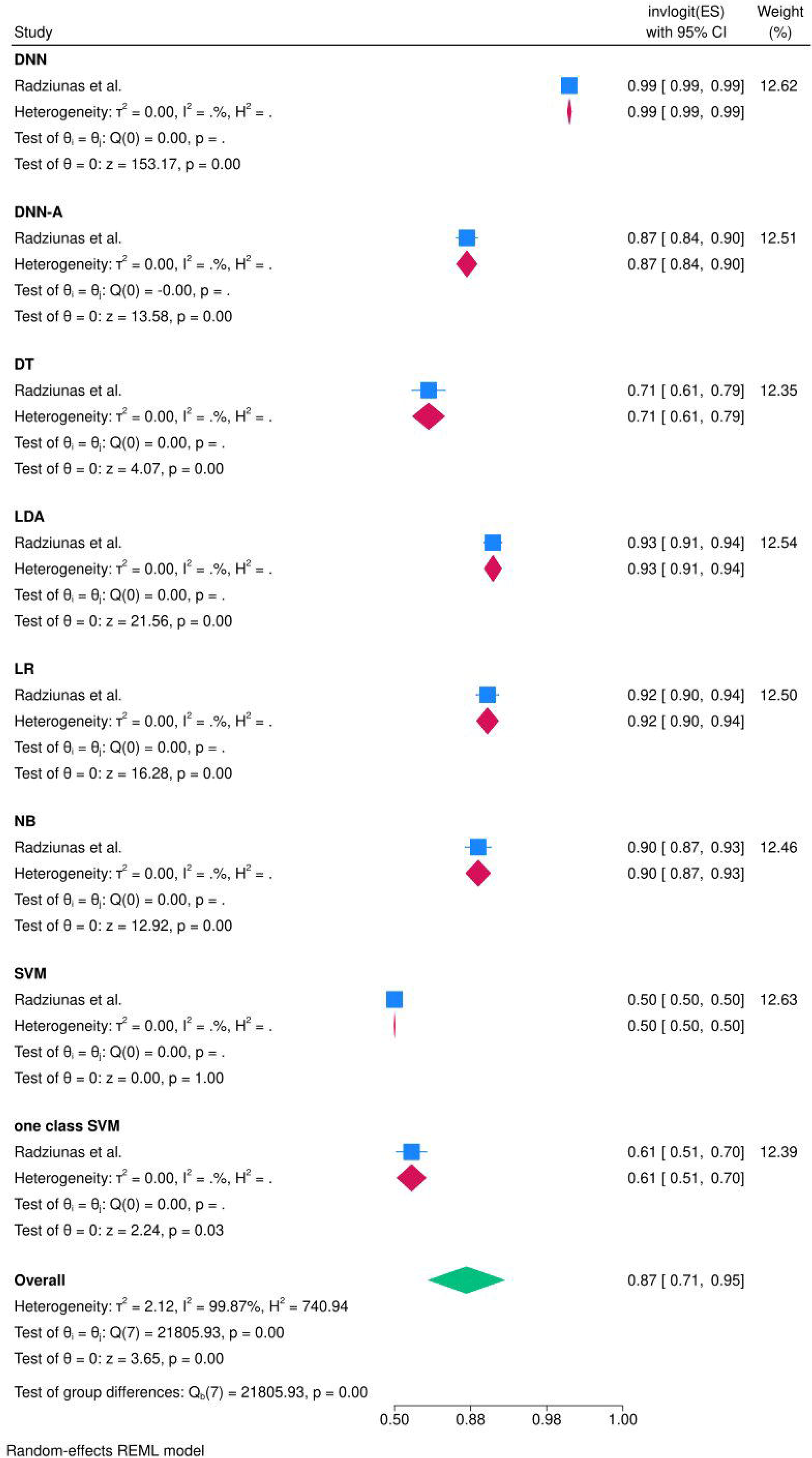
Meta-analysis of AUC of models predicting postoperative delirium.

#### 3.3.3. Other Outcomes

One study developed SVM models based on subcortical volumes or cortical thickness for predicting improvements in dystonia rating scales, which had sensitivities between 50% ^25^ and 79% ^25^. One study built LR, SVM, and RF models based on structural connectivity alone or in combination with demographics for predicting freezing of gait, with AUCs ranging from 65% to 78% ^30^.

### 3.4. Quality Assessment

Regarding the risk of bias assessment with PROBAST+AI, seven studies were classified as low risk, while thirteen were identified as high risk. The high risk of bias was predominantly driven by the analysis domain, which showed concerns across nearly all included studies. In contrast, all studies demonstrated high clinical relevance, with the applicability domain being rated as low concern throughout (Figure 4). Detailed risk of bias assessment using a modified PROBAST+AI approach is provided in Supplemental Table 2.

**Figure 4:**
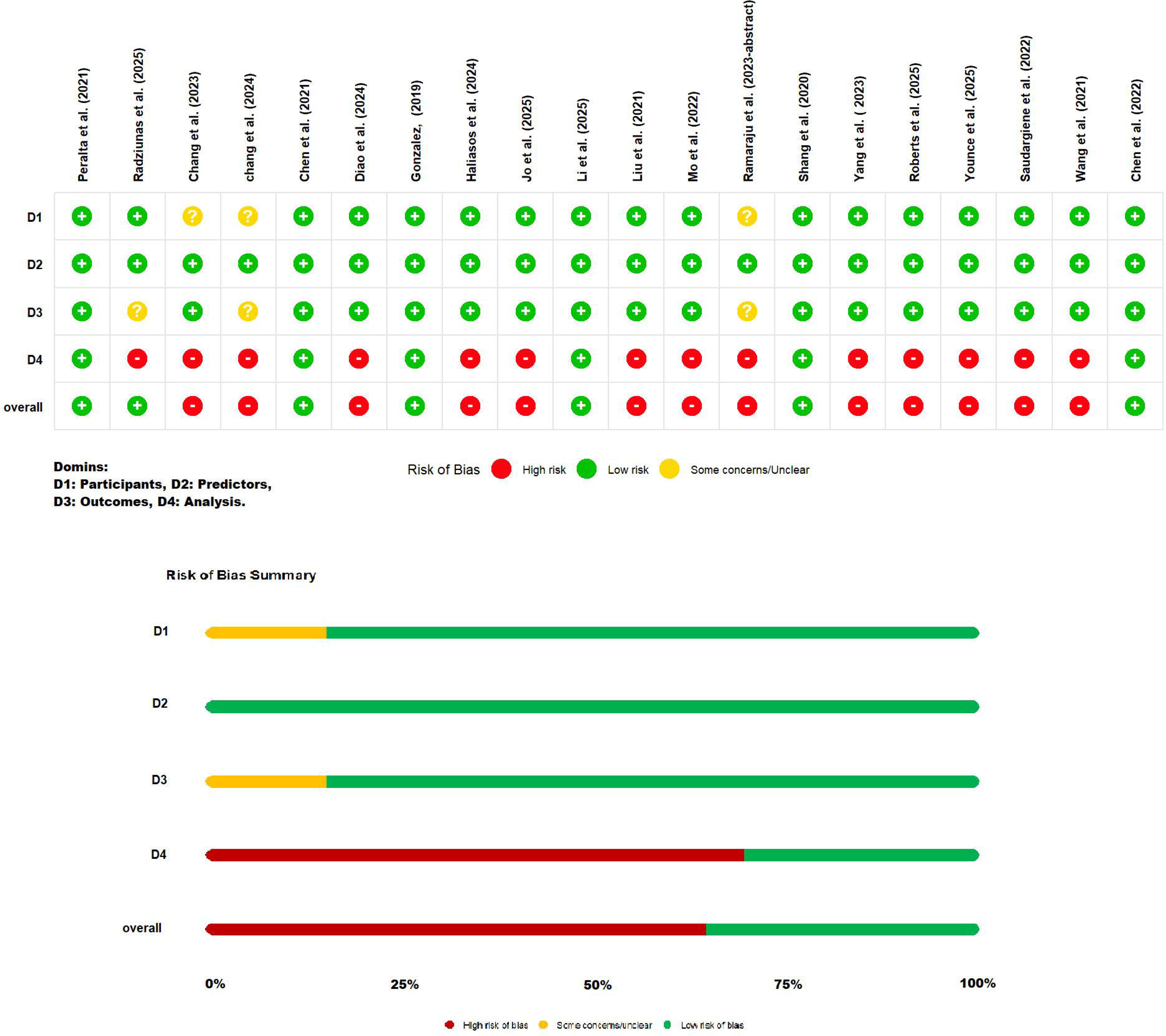
Quality assessment by updated PROBAST+AI

## 4. Discussion

This systematic review and meta-analysis suggest that ML models based on neuroimaging data have potential for predicting postoperative outcomes after DBS in movement disorders. Across 20 studies including 1,551 patients and 53 developed predicting models, most investigations focused on PD, STN-DBS, and postoperative motor improvement measured by UPDRS III, while a smaller number assessed neuropsychiatric outcomes such as delirium, apathy, anxiety, depression, cognition, or freezing of gait. Overall, the pooled discrimination for models predicting motor outcomes was good, with an AUC of 0.86, and the pooled delirium models also showed an AUC of 0.87. These findings support the concept that preoperative neuroimaging contains clinically meaningful information that may help characterize the biological substrate of DBS responsiveness before surgery.

The overall pattern of evidence indicates that imaging-derived biomarkers, particularly radiomics and connectivity-based features, can capture disease-related structural and functional brain variation relevant to DBS outcome. Radiomics was the most frequently used feature set, and many of the highest reported classification performances came from radiomic models. This likely reflects the ability of radiomic pipelines to quantify subtle image heterogeneity not appreciable on routine visual inspection. At the same time, connectivity-based approaches also demonstrated a predictive signal. For example, functional-connectivity models reported MSE values as low as 0.173 and correlation coefficients up to 0.37, while models incorporating structural and functional connectivity achieved an R² of 0.2478. Network-derived structural covariance and structural-connectivity models also showed clinically relevant performance, with AUCs ranging from 65% to 80% and R² values up to 0.67. This is biologically plausible because DBS does not act only at the stimulation target but modulates wider cortico-basal ganglia-thalamic circuits ^38^.

Therefore, models incorporating connectivity-based features, such as functional connectivity, structural connectivity, or structural covariance networks, may provide complementary information to local structural or radiomic features, particularly for outcomes that plausibly depend on distributed circuit function, such as motor improvement, freezing of gait, or neuropsychiatric changes.

Most included studies concentrated on motor outcomes, and this dominance shaped the overall evidence base. The pooled motor AUC of 0.86 suggests that neuroimaging-based ML models can achieve clinically relevant discrimination, but this estimate must be interpreted cautiously. The extremely high heterogeneity indicates that the pooled effect summarizes very different models rather than a single stable effect size. Studies differed markedly in imaging modality, feature engineering, algorithm choice, outcome definition, follow-up duration, and validation approach. Accordingly, the pooled AUC should not be interpreted as the expected performance of a typical model in real-world practice, but rather as evidence that a useful predictive signal exists across multiple neuroimaging domains. The same caution applies to the delirium analysis. Although the pooled AUC was good, it was based on very limited evidence and showed high heterogeneity, so it should be regarded as hypothesis-generating rather than definitive.

An important finding of this study is that model performance varied widely across studies. Some models reported excellent discrimination, with AUCs approaching 0.98 to 0.99, whereas others showed only modest or even poor performance. This variability likely reflects both true differences in modeling quality and differences in the difficulty of the prediction targets. For example, outcomes defined with liberal thresholds or dichotomized in ways that simplify classification may produce inflated discrimination. Similarly, highly selected responder vs non-responder groups can artificially enhance separability. In contrast, studies predicting more complex and clinically heterogeneous outcomes, or using continuous regression targets over longer follow-up intervals, may show lower but more realistic performance. Therefore, the highest reported AUCs should not automatically be viewed as the most clinically credible results.

The quality assessment provides an essential context for interpreting these results. Seven studies were judged as low risk of bias, while thirteen were classified as high risk. Notably, the main source of concern was the analysis domain, whereas the participants, predictors, and outcome domains were generally acceptable, and applicability concerns were low across all included studies. This pattern is important. It suggests that the central limitation of the current literature is not that the wrong patients were studied, the wrong predictors were measured, or the outcomes were clinically irrelevant. Rather, the main problem is how the models were developed, validated, and reported. In other words, the field appears clinically well-directed but methodologically immature.

The predominance of concerns in the analysis domain was driven by recurring issues across studies; small sample sizes, high-dimensional feature spaces, lack of independent external validation, frequent reliance on internal cross-validation alone, occasional data leakage through feature selection performed before resampling, inadequate reporting of calibration, unclear handling of missing data, and the use of complex models without sufficient events per variable. These problems are especially important in neuroimaging-based machine learning, where the number of extracted features often greatly exceeds the number of patients. Under such conditions, apparently excellent performance may reflect overfitting rather than a true generalizable signal. This concern is reinforced by the fact that several studies with very high reported discrimination were still judged high risk because their validation strategy was insufficient to establish robustness.

The contrast between low applicability concerns and high analytical risk is also clinically meaningful. The included cohorts generally represented typical DBS populations, especially patients with PD undergoing STN-DBS, and predictors were usually derived from standardized preoperative imaging pipelines. Outcomes such as UPDRS III improvement were also clinically relevant and familiar to practicing DBS teams. Thus, these studies are asking clinically useful questions in appropriate populations. However, because the models are not yet methodologically secure, translation into routine decision-making remains premature. Put simply, the literature appears closer to proof-of-concept than to implementation.

These findings are broadly consistent with previous reviews on ML and imaging-guided DBS, but this study extends them in several important ways. Earlier reviews described the rapid expansion of ML across the DBS workflow, including candidate selection, surgical planning, electrophysiological signal analysis, adaptive stimulation, programming, and outcome prediction. Peralta et al. reviewed 73 studies of ML in DBS and similarly concluded that the field is methodologically heterogeneous, dominated by classification tasks, and limited by variable validation strategies and reporting standards ^39^.

Watts et al. also emphasized the potential role of ML in PD-DBS while noting that many applications remained preliminary and required stronger prospective and external validation ^40^.

Compared with these broader reviews, the present study focuses more narrowly on preoperative neuroimaging-based ML models for postoperative outcome prediction in movement disorders and adds quantitative synthesis of predictive performance, including pooled AUC estimates for motor and delirium outcomes.

Our results also align with prior Connectomic and imaging-focused DBS literature. Wong et al. reviewed how structural and functional Connectomics can improve DBS targeting and outcome prediction, supporting the concept that DBS response depends on modulation of distributed brain networks rather than stimulation of an isolated anatomical nucleus ^41^.

Similarly, Boutet et al. showed that fMRI responses to DBS could be used with ML to classify optimal versus non-optimal stimulation settings, illustrating the value of network-level imaging biomarkers for individualized DBS therapy ^42^.

The present review complements these studies by showing that such imaging-derived predictive signals are not limited to postoperative programming or contact optimization, but are also detectable preoperatively and may help estimate likely postoperative benefit before surgery.

Moreover, most studies came from China and most focused on PD, with only isolated work in dystonia and other postoperative domains. This narrow distribution limits generalizability. The evidence base currently tells us much more about predicting motor outcome after STN-DBS for PD than about predicting outcomes for GPi-DBS, VIM-DBS, dystonia, tremor, or non-motor endpoints. It also remains uncertain whether models trained in one geographic, technical, or surgical setting would maintain performance across centers with different MRI protocols, surgical workflows, electrode localization methods, or programming strategies. The few studies that included broader or multicenter data are therefore especially valuable, because they move the field closer to clinically transportable models.

The results also suggest that adding clinical or demographic variables to neuroimaging features does not always substantially improve performance. In the limited studies that directly compared multimodal versus imaging-only models, imaging-only approaches were often similar or slightly better. This may indicate that neuroimaging already captures part of the variance represented by conventional clinical markers, or that the clinical features included were too limited to add incremental value. However, this should not be interpreted as evidence against multimodal prediction. Rather, it highlights the need for more carefully designed multimodal studies incorporating standardized clinical variables, medication response, surgical targeting variables, stimulation parameters, and longitudinal follow-up.

Several limitations of this review should be acknowledged. First, substantial heterogeneity across studies limited the extent and interpretability of the meta-analysis. Studies differed in disorder type, DBS target, imaging modality, feature type, algorithm, validation design, follow-up interval, and definition of response. Second, reporting of model performance was inconsistent, with some studies reporting AUC and others MSE, R², correlation coefficients, or only partial classification metrics, reducing comparability. Third, publication bias was significant in the motor outcome meta-analysis, raising the possibility that smaller studies with favorable results were more likely to be published. Fourth, many included studies were small, retrospective, and single-center, which limits confidence in pooled conclusions. Fifth, because the literature is dominated by PD motor outcomes, the conclusions are less secure for dystonia, neuropsychiatric endpoints, and gait-related outcomes. Finally, although this review used structured quality assessment and narrative synthesis, interpretation of heterogeneous machine learning studies inevitably involves some subjectivity.

These limitations mirror the main limitations of the primary literature itself. The current evidence base is constrained by underpowered datasets, limited external validation, insufficient calibration reporting, and a tendency to prioritize discrimination metrics over real-world clinical utility. Few studies evaluated whether predictions would actually improve patient selection, counseling, or treatment planning compared with existing approaches such as clinical assessment and levodopa responsiveness. Likewise, outcome definitions were not uniform, and some studies used thresholds that may not reflect meaningful clinical benefit. Without harmonized endpoints and transparent reporting, it remains difficult to determine whether one modeling strategy is truly superior to another.

Future research should therefore shift from proof-of-concept modeling toward robust translational validation. Large prospective multicenter datasets are needed to improve sample size, capture population diversity, and allow truly independent external validation. Standardization of imaging acquisition, preprocessing, feature extraction, and outcome definitions would reduce heterogeneity and improve comparability. Future studies should also report calibration, decision-curve analysis, missing-data handling, and model interpretability alongside discrimination metrics. Importantly, investigators should avoid leakage-prone pipelines and ensure that all feature selection and tuning steps are performed strictly within resampling frameworks. Multimodal models integrating neuroimaging with clinical, electrophysiological, surgical, and programming variables may provide the most clinically useful predictions, but they must be developed transparently and tested across institutions. Finally, more work is needed on underrepresented indications and outcomes, including dystonia, tremor, cognitive and psychiatric sequelae, gait outcomes, and target-specific predictions for GPi and VIM DBS.

## 5. Conclusion

In conclusion, machine learning models using neuroimaging data show promising potential for preoperative prediction of DBS outcomes, particularly for motor improvement in Parkinson’s disease. However, the current literature is characterized by a persistent gap between encouraging discrimination and reliable clinical readiness. The main weakness of the field lies not in clinical relevance, but in analytical rigor and generalizability. Until larger, externally validated, and methodologically standardized studies are available, these models should be viewed as promising research tools rather than established instruments for routine clinical decision-making.

## Supporting information

Characteristics of the developed AI models

Risk of bias assessment with PROBAST+AI tool

Characteristics of the included studies

## Data Availability

All data produced in the present work are contained in the manuscript

## Acknowledgement

None

## 6. Author’s role

Mohammad-Jasim Golzarian: Conceptualization, Project administration, Investigation, Supervision, Writing – review & editing

Shahryar Rajai Firouzabadi: Investigation, Methodology, Formal analysis, Data curation, Writing – original draft, Writing – review & editing

Zanyar Hajiesmalpoor: Investigation, Validation, Writing – original draft Zahra Azizan: Investigation, Writing – original draft

Alireza Alikhani: Visualization, Investigation, Writing – original draft Pourya Moshayedi: Validation, Writing – review & editing

## 9. Ethical compliance statement

We confirm that we have read the Journal’s position on issues involved in ethical publication and affirm that this article is consistent with those guidelines. The present paper is also approved by ethics committee of Shahid Beheshti University of medical sciences. Informed patient consent was not necessary for this article.

## 10. Legends for Supplemental files

Supplemental Table 1: Characteristics of the developed AI models Supplemental Table 2: Risk of bias assessment with PROBAST+AI tool

## 11. Legend for Table

Table 1: Characteristics of the included studies

---

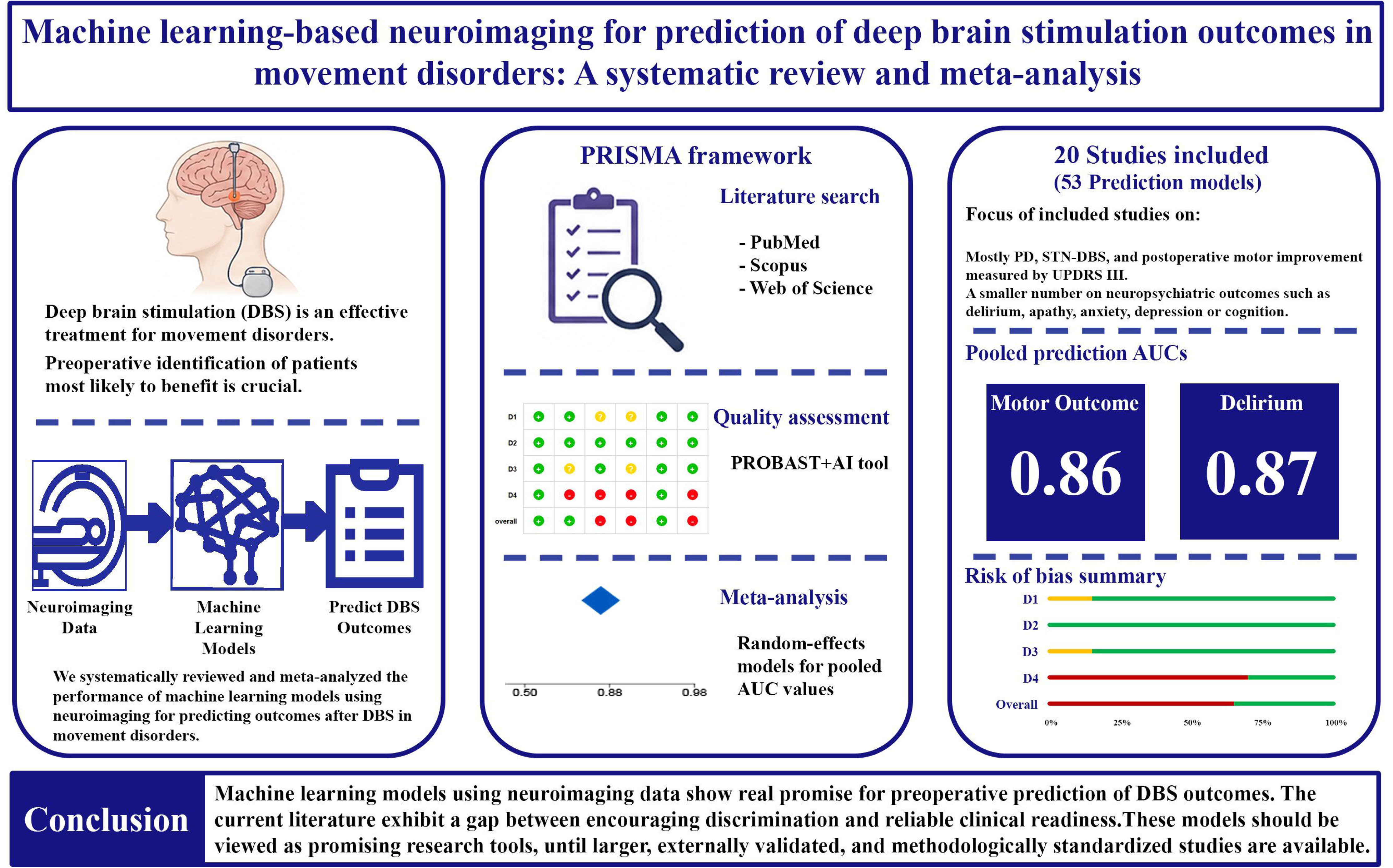

