## Supplementary material for "Machine learning-based neuroimaging for prediction of deep brain stimulation outcomes in movement disorders: Systematic review and meta-analysis": Characteristics of the developed AI models

Supplemental Table 1: Characteristics of the developed AI models.

| Author, Year  Country | Algorithm used | Features | Validation | Performance metrics | | | | | |
| --- | --- | --- | --- | --- | --- | --- | --- | --- | --- |
|  |  |  |  | Sensitivity | Specificity | AUC | MSE | MAE | R^2^ |
| Radziunas et al., 2025  Lithuania | **DNN** | Radiomics | K-fold cross-validation | **100%** | **99%** | **99%** |  |  |  |
|  | LR |  |  | 92% | 92% | 92% |  |  |  |
|  | DT |  |  | 55% | 87% | 71% |  |  |  |
|  | LDA |  |  | 96% | 91% | 93% |  |  |  |
|  | NB |  |  | 82% | 99% | 90% |  |  |  |
|  | SVM |  |  | 0% | 100% | 50% |  |  |  |
|  | one class SVM |  |  | 89% | 33% | 61% |  |  |  |
|  | DNN-A |  |  | 92% | 82% | 87% |  |  |  |
| Chang et al., 2023  China | **SVR** | **Serum UA related functional connectivity** | Leave-one out cross-validation |  |  |  | **0.173** |  |  |
| Chang et al., 2024  China | **CNN** | **Radiomics + multi-instance learning** | K-fold cross-validation | **80%** | **75%** | **87%** |  |  |  |
| Chen et al., 2021  China | **SVM** | **Brain morphology** | Train-test |  |  |  |  |  | **0.3224** |
| Chen et al., 2022  China | **SVM** | **Structural and functional connectivity** | Train-test |  |  |  |  |  | **0.2478** |
| Diao et al., 2024  China | **XGBoost** | **SCN** | Train-test |  |  | **80%** | **0.06** |  | **0.67** |
| Gonzalez-Escamilla et al., 2019  Germany | **SVM** | **Subcortical volumes** | Train-test | **79%** |  |  |  |  |  |
|  | SVM | Cortical thickness of regions selected using a SCN |  | **50%** |  |  |  |  |  |
| Haliasos et al., 2024  UK | **RF** | **Radiomics** | Train-test with bootstrapping | **80%** | **90%** | **92%** |  |  |  |
|  | LR |  |  | 75% | 80% | 86% |  |  |  |
|  | SVM |  |  | 72% | 81% | 86% |  |  |  |
|  | NB |  |  | 78% | 86% | 90% |  |  |  |
|  | KNN |  |  | 71% | 80% | 83% |  |  |  |
| Jo et al., 2025  South Korea | **LR** | **Structural Connectivity** | K-fold cross-validation | **71%** | **79%** | **78%** |  |  |  |
|  |  | Demographics and structural connectivity |  | 71% | 79% | 74% |  |  |  |
|  | SVM | Structural Connectivity |  | 76% | 79% | 75% |  |  |  |
|  |  | Demographics and structural connectivity |  | 71% | 79% | 75% |  |  |  |
|  | RF | Structural Connectivity |  | 47% | 68% | 65% |  |  |  |
|  |  | Demographics and structural connectivity |  | 47% | 63% | 66% |  |  |  |
| Li et al., 2025  China | LR | Radiomics | Train-test | 83% | 86% | 84% |  |  |  |
|  | SVM |  |  | 78% | 93% | 85% |  |  |  |
|  | **RF** |  |  | **83%** | **88%** | **86%** |  |  |  |
| Liu et al., 2021  China | **LR** | **Radiomics** | Leave-one out cross-validation | **80%** | **85%** | **85%** |  |  |  |
|  |  | Radiomics/LCT |  | 73% | 77% | 83% |  |  |  |
| Peralta et al., 2021  France | SVM | Preoperative clinical scores extracted via ANN | K-fold cross-validation |  |  |  |  |  |  |
| Mo et al., 2022  China | **SVM** | Surface based morphometry | Leave-one out cross-validation |  |  |  |  |  | **0.14** |
| Ramaraju et al., 2023  India | **LR** | Radiomics | NP |  |  | **98%** |  |  |  |
|  | DT |  |  |  |  |  |  |  |  |
|  | NB |  |  |  |  |  |  |  |  |
|  | SVM |  |  |  |  |  |  |  |  |
|  | DNN |  |  |  |  | 87% |  |  |  |
| Roberts et al., 2024  USA | **LASSO regression** | Radiomics | Nested cross-validation |  |  | **99%** |  |  |  |
| Saudargiene et al., 2022  Lithuania | **LR** | Radiomics | Train-test with bootstrapping | **99%** | **96%** | **98%** |  |  |  |
|  | DT |  |  | 15% | 90% | 52% |  |  |  |
|  | LDA |  |  | 83% | 77% | 80% |  |  |  |
|  | NB |  |  | 22% | 99% | 61% |  |  |  |
|  | SVM |  |  | 60% | 98% | 79% |  |  |  |
|  | DNN |  |  | 87% | 87% | 87% |  |  |  |
|  | one class SVM |  |  | 24% | 71% | 48% |  |  |  |
|  | DNN-A |  |  | 73% | 68% | 71% |  |  |  |
| Shang et al., 2020  China | SVR | Functional connectivity | Nested cross-validation |  |  |  |  |  | 398.57 |
|  | **Gradient boosting machine** |  |  |  |  |  |  |  | **240.74** |
|  | RF |  |  |  |  |  |  |  | 282.13 |
|  | Linear regression |  |  |  |  |  |  |  | 862.94 |
|  | Ridge regression |  |  |  |  |  |  |  | 573.95 |
|  | LASSO regression |  |  |  |  |  |  |  | 411.83 |
| Wang et al., 2021  USA | **Ridge regression** | Functional connectivity based on hemispheric asymmetry selected by RF | Nested cross-validation |  |  |  |  |  |  |
| Yang et al., 2023  China | **CatBoost** | Amplitudes of low frequency fluctuations | K-fold cross-validation | **80%** | **93%** | **94%** |  |  |  |
|  | Ensemble model^*^ |  |  |  |  |  |  | 0.10 |  |
| Younce et al., 2025  USA | **LASSO regression** | Functional connectivity + clinical + brain morphology | Leave-one out cross-validation |  |  |  |  |  | **0.3153** |

^Bold values represent the best performing model.
*: ensemble of linear regression/ridge regression/k-adjacency regression/AdaBoost/Gradient boosting/SVM)
abbreviations: AUC: Area Under Receiver Operating Curve/ MSE: Mean Standard Error/ MAE: Mean Absolute Error/ DNN: Deep neural Network/ LR: Logistic Regression/ DT: Decision Tree/ LDA: Linear Discriminant Analysis/ NB: Naïve Bayesian/ SVM: Support Vector Machine/ SVR: Support Vector Regression/ DNN-A: Deep Neural network-Autoencoder/ CNN: Convolutional Neural Network/ RF: Random Forest/ KNN: K-Nearest Neighbor/ UA: Uric Acid/ SCN: Structural Covariance Network/ LCT: Levodopa Challenge Test/ ANN: Artificial Neural Network/ NP: Not Provided/^
