## Supplementary material for "Machine learning-based neuroimaging for prediction of deep brain stimulation outcomes in movement disorders: Systematic review and meta-analysis": Risk of bias assessment with PROBAST+AI tool

| 1- **Radziunas et al. (2025)** | | | | |
| --- | --- | --- | --- | --- |
| Domain | Risk of bias (Low/High/Unclear) | Applicability (Low/High/Unclear | Justification (1-2 sentences) | Page/section in PDF |
| Participants | Low | Low | Inclusion/exclusion criteria were clearly defined based on CAPSIT-PD guidelines, and the sample represents typical PD candidates for STN-DBS. | Page 3,5, Methods/ procedure |
| Predictors | Low | Low | Radiomic features were extracted from preoperative T1W and T2W MRI using standardized pipelines (FreeSurfer/PyRadiomics) without knowledge of the outcome. | Page 11, Image Segmentation |
| Outcome | Probably Low | Low | Postoperative delirium was assessed using the 4AT scale, which is a validated and objective tool in clinical settings.  There was no information about the blinding. | Page 5, Procedures |
| Analysis | High |  | While cross-validation was used, the sample size is very small (n=34) with only 7 positive cases, and no independent external validation was performed, increasing the risk of overfitting. | Page 12, Results/Discussion |
| Overall PROBAST | High | Low | The study is classified as High Risk due to the small sample size and lack of external validation in the machine learning analysis (Domain 4). | Final Judgment |

**Supplemental Table 2:** Risk of bias assessment with PROBAST+AI tool

| 2- **Chang et al. (2023)** | | | | |
| --- | --- | --- | --- | --- |
| Domain | Risk of bias (Low/High/Unclear) | Applicability (Low/High/Unclear | Justification (1-2 sentences) | Page/section in PDF |
| Participants | Probably Low / unclear / yellow | Low | Inclusion and exclusion criteria were clearly stated; patients had intermediate-to-advanced PD, which is the standard population for DBS. Single center and small sample size | Page 2, Section 2.1 |
| Predictors | Low | Low | Serum Uric Acid (UA) was measured 5 days before surgery, and rs-fMRI was performed in the OFF-medication state before surgery, ensuring predictors were measured before the outcome. | Page 3, Section 2.2 & 2.3 |
| Outcome | Low | Low | The outcome was the UPDRS-III improvement rate at 2 years post-op, which is a standard and reliable clinical measure for DBS efficacy. | Page 3, Section 2.2 |
| Analysis | High |  | The sample size is small (n=32), and the study used Leave-One-Out Cross-Validation (LOOCV) without any external validation, which highly increases the risk of overfitting in SVR models. | Page 4, Section 2.8 / Page 8, Discussion |
| Overall PROBAST | High | Low | The study is rated as High Risk overall because of the statistical limitations in the machine learning analysis (small n and lack of external validation) | Final Judgment |

| 3- **Chang et al. (2024)** | | | | |
| --- | --- | --- | --- | --- |
| Domain | Risk of bias (Low/High/Unclear) | Applicability (Low/High/Unclear | Justification (1-2 sentences) | Page/section in PDF |
| Participants | Probably Low / yellow | Low | A relatively large cohort (n=127) was used with clear inclusion/exclusion criteria. The population is representative of advanced PD patients. Single center and small sample size | Page 2, Patients |
| Predictors | Low | Low | T1-weighted MRI scans were acquired 3 days before surgery, ensuring predictors were measured before the clinical outcome. | Page 2, Patients and Image segmentation |
| Outcome | Probably Low/ yellow | Low | Outcomes were defined based on the UPDRS-III improvement rate at 2 years, which is a standard and robust clinical endpoint. the cut off point for improvement is questionable. | Page 3, Patients |
| Analysis | High |  | Although 5-fold cross-validation and SMOTE were used, the study lacks independent external validation, and the categorization of "relief" (improvement > 0) is very liberal, potentially introducing bias. Also, the usage of 3D model necessitates larger sample size, though mitigate the risk of overfitting. | Page 4, Signature Building , multi-instance learning |
| Overall PROBAST | High | Low | The overall risk is High primarily due to the lack of external validation and the choice of a low threshold for the positive outcome (improvement > 0). | Final Judgment |

| 4- **Chen et al. (2021)** | | | | |
| --- | --- | --- | --- | --- |
| Domain | Risk of bias (Low/High/Unclear) | Applicability (Low/High/Unclear | Justification (1-2 sentences) | Page/section in PDF |
| Participants | Low | Low | 94 PD patients were retrospectively enrolled with clear diagnostic criteria (UK Brain Bank) and balanced training/test sets. | Page 668, Patients |
| Predictors | Low | Low | Brain morphology (cortical thickness) and VTA were measured using standardized MRI/CT protocols before evaluating the stimulation response. | Page 668-669, Neuroimage |
| Outcome | Low | Low | The initial stimulation response was objectively measured using the MDS-UPDRS III scale 4–5 weeks after surgery. | Page 668, Evaluations |
| Analysis | Probably low |  | Although feature selection (LASSO) and SVM were used, the small sample size (n=73 for training, n=21 for testing) is the important weakness. the study lacks independent external validation. Statistical analyses were only performed within the training set, avoiding information leakage. | Page 669, Prediction |
| Overall PROBAST | Low | Low | The study is rated as High Risk due to the lack of an external validation cohort, which is a requirement for Low Risk in PROBAST-AI. | Final Judgment |

| 5- **Diao et al. (2024)** | | | | |
| --- | --- | --- | --- | --- |
| Domain | Risk of bias (Low/High/Unclear) | Applicability (Low/High/Unclear | Justification (1-2 sentences) | Page/section in PDF |
| Participants | Low | Low | A large sample of 138 PD patients and 40 HCs was used with clear inclusion/exclusion and long-term follow-up (>1 year). | Page 1107, Participants |
| Predictors | Low | Low | Preoperative T1 MRI was used to construct individual networks (ISCN) through a standardized and objective pipeline (NTP). | Page 1108, ISCN Measures |
| Outcome | Low | Low | Long-term motor improvement (MDS-UPDRS III) was assessed 1–3 years postoperatively, reflecting a stable treatment effect. | Page 1108, Clinical Exam |
| Analysis | High |  | Advanced ML (XGBoost and MLP) was used with a 75:25 split, but the study lacks independent external validation to ensure model generalizability. overfitting and insufficient validation. selection for machine learning models, use of extreme responder groups only, and absence of pre-specified analysis plan | Page 1109, Prediction |
| Overall PROBAST | High | Low | Rated as High Risk due to the reliance on internal validation (train/test split) without an external cohort, as per PROBAST-AI standards. | Final Judgment |

| 6- **Gonzalez, 2019** | | | | |
| --- | --- | --- | --- | --- |
| Domain | Risk of bias (Low/High/Unclear) | Applicability (Low/High/Unclear | Justification (1-2 sentences) | Page/section in PDF |
| Participants | Low | Low | The study included a well-defined multicenter cohort of primary dystonia patients with clear inclusion criteria and handled poor image quality appropriately to avoid selection bias. | Page 2 / Methods (Study Participants) |
| Predictors | Low | Low | Predictors were derived from standard preoperative MRI using validated automated pipelines (FreeSurfer) and were assessed blinded to the clinical outcome. | Page 3 / Methods (Cortical Thickness Maps) |
| Outcome | Low | Low | Outcome was measured using standardized clinical scales (BFMDRS/TWSTRS) at a stable 3-year follow-up, with clinicians blinded to the study's imaging data. | Page 2 / Methods (GPi-DBS Clinical Outcomes) |
| Analysis | Low | - | The study used SVM with 10-fold cross-validation and, crucially, performed external validation on an independent cohort, which effectively mitigated overfitting risks. | Page 3-4 / Methods (GPi-DBS Outcome-Based Classification) |
| Overall PROBAST | Low | Low | The study demonstrates high methodological rigor, especially through its use of an independent test cohort for model validation. | Final judgment |

| 7- **Haliasos et al., 2024** | | | | |
| --- | --- | --- | --- | --- |
| Domain | Risk of bias (Low/High/Unclear) | Applicability (Low/High/Unclear | Justification (1-2 sentences) | Page/section in PDF |
| Participants | Low | Low | The study used a retrospective cohort with clear inclusion and exclusion criteria, representing typical Parkinson's disease candidates for STN-DBS | Page 3 / Materials and Methods (Patient selection) |
| Predictors | Low | Low | Predictors were derived from standard preoperative T1-weighted MRI using automated radiomics extraction (PyRadiomics), ensuring objective and reproducible feature measurement. | Page 4-5 / Materials and Methods (MRI acquisition / Image processing) |
| Outcome | Low | Low | The outcome was based on a clinically meaningful 5-point improvement in the standardized UPDRS-III motor score at a 1-year follow-up. | Page 3-4 / Materials and Methods (Patient selection) |
| Analysis | High |  | The study lacks independent external validation, and the exceptionally high performance (AUC 0.99) on a relatively small dataset suggests a significant risk of overfitting. Also, feature selection was performed on the entire dataset before resampling, leading to potential data leakage and overfitting. | Page 5 / Machine learning models; Page 11 / Study limitations |
| Overall PROBAST | High | Low | Although the study is a strong proof-of-concept, the overall risk of bias is high due to the lack of external validation for the machine learning models. | Final Judgment |

| 8- **Jo et al., 2025** | | | | |
| --- | --- | --- | --- | --- |
| Domain | Risk of bias (Low/High/Unclear) | Applicability (Low/High/Unclear | Justification (1-2 sentences) | Page/section in PDF |
| Participants | Low | Low | The study utilized a well-characterized retrospective cohort of PD patients with specific "off-freezing" symptoms, ensuring clinical relevance for GPi-DBS candidates. | Page 2 / Materials & Methods (Study population) |
| Predictors | Low | Low | Predictors were derived from preoperative DTI and VAT-based structural connectivity using the validated Lead-DBS pipeline, which is a standard approach in connectomic studies. | Page 3 / Materials & Methods (Image acquisition / Electrode localization) |
| Outcome | Low | Low | The outcome was assessed using the MDS-UPDRS Part III (Item 11) with excellent inter-rater reliability (ICC 0.97) under standardized medication/stimulation conditions. | Page 2-3 / Materials & Methods (Surgery and evaluation) |
| Analysis | High | - | While 10-fold cross-validation was used, the small sample size (n=58) increases the risk of overfitting in machine learning models. the study did not check if the model's predictions were well-calibrated, did not explain how missing data was handle. | Page 4 / Statistical analysis; Page 8 / Discussion (Limitations) |
| Overall PROBAST | High | Low | Despite the high clinical value and rigorous neuroimaging pipeline, the overall risk of bias is high due to the lack of external validation and a relatively small dataset. | Final judgment |

| 9- **Li et al., 2025** | | | | |
| --- | --- | --- | --- | --- |
| Domain | Risk of bias (Low/High/Unclear) | Applicability (Low/High/Unclear | Justification (1-2 sentences) | Page/section in PDF |
| Participants | Low | Low | The study enrolled a substantial cohort of 209 PD patients with clear criteria for "Good" vs. "Poor" improvement, reflecting a realistic clinical spectrum for STN-DBS | Page 2-3 / Methods (Patients) |
| Predictors | Low | Low | Predictors were objective radiomics features extracted from preoperative T2-MRI using standardized tools (PyRadiomics), with high inter-observer agreement (ICC > 0.75). | Page 3-4 / Methods (ROI segmentation and features extraction) |
| Outcome | Low | Low | Improvement was defined using the standardized MDS-UPDRS III score at a 3-month follow-up, which is a standard and clinically relevant metric for motor function. | Page 3 / Methods (Clinical data assessments) |
| Analysis | Low | - | the study appropriately split the data into training and test sets, applied feature selection only on the training set, and validated model performance using ROC, calibration, and decision curve analysis, minimizing the risk of overfitting. | Page 4-5 / Methods (Selection of radiomics feature, Construction of ML models); Page 10 / Discussion (Limitations) |
| Overall PROBAST | Low | Low | Clear participant selection, standardized outcome definition, robust predictor handling, and rigorous model validation collectively support a low risk of bias for the study. | Final judgment |

| 10- **Liu et al., 2021** | | | | |
| --- | --- | --- | --- | --- |
| Domain | Risk of bias (Low/High/Unclear) | Applicability (Low/High/Unclear | Justification (1-2 sentences) | Page/section in PDF |
| Participants | Low | Low | The study prospectively recruited 33 PD patients with clear inclusion/exclusion criteria, ensuring the cohort is representative of DBS candidates. | Page 3 / Materials and Methods (Patients) |
| Predictors | Low | Low | Predictors were objective radiomics features extracted from preoperative QSM images using an automated pipeline (PyRadiomics), with a strong focus on inter-rater stability (ICC > 0.85). | Page 4-5 / Materials and Methods (Image Pre-processing / Feature Extraction) |
| Outcome | Low | Low | The outcome was based on the standard MDS-UPDRS III motor score improvement with a clear 30% cutoff at a 6-month follow-up. | Page 3 / Materials and Methods (Clinical Evaluation) |
| Analysis | High | - | The sample size is very small (n=33) and the study relied on Leave-One-Out Cross-Validation (LOOCV) without any independent external validation, which carries a high risk of overfitting. | Page 5 / Development and Validation; Page 9 / Discussion (Limitations) |
| Overall PROBAST | High | Low | While the use of QSM radiomics is innovative, the overall risk of bias is high due to the small pilot nature of the study and the lack of external validation for its predictive models. | Final judgment |

| 11- **Mo et al., 2022** | | | | |
| --- | --- | --- | --- | --- |
| Domain | Risk of bias (Low/High/Unclear) | Applicability (Low/High/Unclear | Justification (1-2 sentences) | Page/section in PDF |
| Participants | Low | Low | The study included 78 PD patients and 55 healthy controls with well-documented clinical assessments, making the cohort representative of the PD population | Page 2-3 / Methods (Participants) |
| Predictors | Low | Low | Predictors are objective surface-based morphological metrics (thickness, sulcal depth, etc.) estimated using the validated CAT12 automated pipeline. | Page 3 / Methods (Computation of surface-based morphometry) |
| Outcome | low | Low | Treatment response was evaluated using standard MDS-UPDRS III scores for both medication and DBS, which are gold-standard metrics in PD research. | Page 3 / Methods (Participants) |
| Analysis | High | - | The machine learning part uses Leave-One-Out Cross-Validation (LOOCV) on a limited sample without independent external validation, which is a major source of potential overfitting. Also, feature selection was performed on the full dataset prior to cross-validation, leading to data leakage and an increased risk of overfitting. | Page 4 / Methods (Prediction model); Page 9 / Discussion (Limitations) |
| Overall PROBAST | High | Low | Although the study provides a comprehensive cortical analysis, the lack of external validation for its predictive models results in a high risk of bias according to PROBAST-AI. | Final Judgment |

| 12- **Ramaraju et al (2023-abstract)** | | | | |
| --- | --- | --- | --- | --- |
| Domain | Risk of bias (Low/High/Unclear) | Applicability (Low/High/Unclear | Justification (1-2 sentences) | Page/section in PDF |
| Participants | Unclear | Low | The study included 34 Parkinson’s disease subjects undergoing STN-DBS, which is a representative clinical population for this intervention. | Section 121797 / Methods |
| Predictors | Low | Low | Predictors are objective radiomic textures (120 features per nucleus) extracted from pre-op MRI using the standardized Py-Radiomics and FreeSurfer pipelines. | Section 121797 / Methods |
| Outcome | Unclear | Low | The outcome (motor response) was assessed postoperatively, with a clear distinction between poor, good, and very-good responses. | Section 121797 / Methods & Results |
| Analysis | High / unclear |  | With only 34 subjects and 5,040 feature manifestations, the study faces a severe "p >> n" problem; despite using mRMR for feature reduction, the exceptionally high accuracy (96.65%) strongly suggests overfitting without external validation. |  |
| Overall PROBAST | High | Low | Although the study uses advanced AI techniques and high-quality pipelines, the small sample size and lack of independent validation result in a high risk of bias. | Final Judgment |

| 13- **Shang et al., 2020** | | | | |
| --- | --- | --- | --- | --- |
| Domain | Risk of bias (Low/High/Unclear) | Applicability (Low/High/Unclear | Justification (1-2 sentences) | Page/section in PDF |
| Participants | Low | Low | The study included 50 PD patients suitable for STN-DBS based on standardized levodopa challenge tests, representing a typical clinical surgical cohort. | Page 2 / Materials and Methods (Participants) |
| Predictors | Low | Low | Predictors were derived from objective whole-brain functional connectivity using the Brainnetome Atlas (246 regions) and preprocessed with standardized pipelines (SPM12). | Page 3 / Image Preprocessing and Brain Network Construction |
| Outcome | Low |  | The outcome was measured as the percentage change in the MDS-UPDRS III score, which is the gold standard for assessing motor improvement in DBS. | Page 2 / Materials and Methods (Participants) |
| Analysis | Low |  | Nested cross-validation minimized data leakage and provided realistic performance estimates, but lack of external validation and small sample size (n = 50) relative to 242 features gives a low-to-moderate risk of bias | Page 3 / Connectome-Based Predictive Modeling; Page 6 / Discussion (Limitations) |
| Overall PROBAST | Low | Low | The study provides high clinical relevance and is classified as "low-moderate Risk of Bias" primarily due to the small sample size and the lack of external validation on an independent dataset. | Final Judgment |

| 14- **Yang et al., 2023** | | | | |
| --- | --- | --- | --- | --- |
| Domain | Risk of bias (Low/High/Unclear) | Applicability (Low/High/Unclear | Justification (1-2 sentences) | Page/section in PDF |
| Participants | Low | Low | The study included 44 PD patients and 44 healthy controls with strict surgical and imaging inclusion/exclusion criteria, ensuring a high-quality clinical sample. | Page 2 / Methods (Participants) |
| Predictors | Low | Low | Predictors were objective ALFF maps derived from rs-fMRI using a standardized and reliable preprocessing pipeline (DPARSF), independent of ROI hypotheses. | Page 2-3 / Methods (MRI data acquisition and post-processing) |
| Outcome | Low | Low | The outcome was measured using the gold-standard MDS-UPDRS III improvement ratio, and responders were categorized with a clear 50% threshold. | Page 2 / Methods (Surgical procedures and clinical evaluation) |
| Analysis | High – moderate |  | Although the study used robust ensemble methods and nested 10-fold cross-validation, the sample size (n=44) is still small for high-dimensional fMRI data, and it lacks external validation on an independent cohort. | Page 3 / Predictive models; Page 5 / Discussion (Strengths and limitations) |
| Overall PROBAST | High | Low | While the study demonstrates an impressive AUC (0.94) and high clinical applicability, it is rated as "High Risk of Bias" due to the common limitation of small sample size and lack of independent external validation. | Final Judgment |

| 15- **Roberts et al., 2025** | | | | |
| --- | --- | --- | --- | --- |
| Domain | Risk of bias (Low/High/Unclear) | Applicability (Low/High/Unclear | Justification (1-2 sentences) | Page/section in PDF |
| Participants | Low | Low | Multi-center study (Center 1 & 2) with 67 patients, using standardized MDS criteria and clear inclusion/exclusion protocols. | Page 2 / Methods (Patient Cohorts) |
| Predictors | Low | Low | Radiomic features were extracted from QSM using the well-validated MEDI-L1 algorithm and PyRadiomics, ensuring objective and high-quality predictors. | Page 2 / Methods (Data Preprocessing) |
| Outcome | Low | Low | Outcome was defined by the UPDRS-III improvement ratio at 6 months, which is a robust and clinically relevant metric for DBS success. | Page 2 / Methods (Data Acquisition) |
| Analysis | High |  | While the study used advanced "Label Noise Compensation" and nested cross-validation to mitigate overfitting, the sample size is still relatively small, though significantly more robust than previous pilot studies. | Page 3 / Regression Model & Training |
| Overall PROBAST | High | Low | This study is a significant step forward; by using multi-center data and novel data augmentation (Noise Injection), it reduces the risk of bias compared to earlier single-center pilot studies. | Final Judgment |

| 16- **Younce et al. (2025)** | | | | |
| --- | --- | --- | --- | --- |
| Domain | Risk of bias (Low/High/Unclear) | Applicability (Low/High/Unclear | Justification (1-2 sentences) | Page/section in PDF |
| Participants | Low | Low | he study used a well-defined cohort of 65 PD patients with clear inclusion/exclusion criteria (UK Brain Bank) and rigorous quality control for DBS candidacy. | Page 2 / Methods (Participants) |
| Predictors | Low | Low | Predictors include pre-operative clinical data, volumetric segmentation (FreeSurfer 7.3), and rs-fcMRI, all obtained through objective and standardized pipelines. | Page 3 / MRI Methods |
| Outcome | Low | Low | The outcome was the relative change in UPDRS-III scores, averaged over 12 months post-DBS to ensure stability and reduce random fluctuations. | Page 2-3 / Clinical Evaluation |
| Analysis | High |  | Relaxed LASSO and LOOCV were used to mitigate overfitting; however, a calibration plot was not provided, and the sample size (n=65) is relatively small for the number of predictors, and it lacks external validation on an independent cohort. | Page 4 / Statistical Analysis; Page 7 / Discussion (Limitations) |
| Overall PROBAST | High | Low | Despite the advanced multimodal approach, the model is at high risk of bias due to the absence of independent external validation, which is common in single-center pilot studies. | Final Judgment |

| 17- **Saudargiene et al., 2022** | | | | |
| --- | --- | --- | --- | --- |
| Domain | Risk of bias (Low/High/Unclear) | Applicability (Low/High/Unclear | Justification (1-2 sentences) | Page/section in PDF |
| Participants | Low | Low | The study included 34 PD patients selected via CAPSIT-PD guidelines, with a well-defined follow-up and clinical profiling. | Page 3 / Study subject |
| Predictors | Low | Low | Objective radiomic features were extracted from pre-op MRI using standardized tools (FreeSurfer 7.0 and PyRadiomics) for amygdala and hippocampus regions. | Page 3 / Image pre-processing and segmentation |
| Outcome | Low | Low | The outcome was based on the PDCS (Parkinson’s Disease Composite Scale) improvement (>30%), which is a valid and objective clinical metric. | Page 3 / Study subjects |
| Analysis | High |  | With only 34 patients and 20 selected features, the sample-to-feature ratio is very low, leading to a high risk of overfitting, especially with the reported 96% accuracy. | Page 4 / Radiomic feature selection & ML models |
| Overall PROBAST | High | Low | Despite the innovative approach and high reported accuracy, the extremely small sample size and lack of independent validation result in a high risk of bias. | Final Judgment |

| 18- **Wang et al., 2021** | | | | |
| --- | --- | --- | --- | --- |
| Domain | Risk of bias (Low/High/Unclear) | Applicability (Low/High/Unclear | Justification (1-2 sentences) | Page/section in PDF |
| Participants | Low | Low | 55 patients with idiopathic PD were included, and the study used clear UK Brain Bank criteria for diagnosis and surgery candidacy. | Page 2 / Methods (Participants) |
| Predictors | Low | Low | Predictors are objective functional connectivity metrics (rs-fMRI) categorized into LH, RH, Ho, and He connections using the Brainnetome Atlas. | Page 3 / Image Preprocessing |
| Outcome | Low | Low | The outcome was the improvement rate of UPDRS-III scores measured 6 months post-DBS, which is a standard clinical measure. | Page 3 / Methods |
| Analysis | High |  | The study used Random Forest for feature selection and Ridge Regression, which helps prevent overfitting, but lacks external validation. | Page 3-4 / Statistical Analysis |
| Overall PROBAST | High | Low | The risk is high because, although they used 55 patients (more than many pilots), the results still rely on a single-center cohort and lack independent testing | Final Judgment |

| 19- **Chen et al., 2022** | | | | |
| --- | --- | --- | --- | --- |
| Domain | Risk of bias (Low/High/Unclear) | Applicability (Low/High/Unclear | Justification (1-2 sentences) | Page/section in PDF |
| Participants | Low | Low | 98 patients were enrolled (78 training / 20 test), which is a solid sample size for a predictive connectivity study. | Page 2 / Study Participant |
| Predictors | Low | Low | Used seed-based functional and structural connectivity from the PPMI database, linked to the patient-specific VTA. | Page 3 / Connectivity Evaluation |
| Outcome | Low | Low | Initial motor outcome (4-5 weeks post-op) was measured using MDS-UPDRS III, providing a consistent short-term metric. | Page 2 / Study Participants |
| Analysis | Low |  | Rigorous ML pipeline using LASSO for feature reduction and SVM with Genetic Algorithm optimization, validated on an independent test set. | Page 3 / Predicting STN-DBS Outcome |
| Overall PROBAST | Low | Low | The study is high-quality and low bias due to its use of a separate test set and well-established normative connectomes for validation. | Final Judgment |

| 20- **Peralta et al. 2021** | | | | |
| --- | --- | --- | --- | --- |
| Domain | Risk of bias (Low/High/Unclear) | Applicability (Low/High/Unclear | Justification (1-2 sentences) | Page/section in PDF |
| Participants | Low | Low | A large cohort of 196 PD patients was used, covering multiple targets (STN, GPi, VIM), which increases generalizability. | Page 2 / Data |
| Predictors | Low | Low | Multimodal: includes pre-op clinical scores (compressed via AE), striatal shape (T1-MRI), and demographics. | Page 3 / Proposed method |
| Outcome | Low | Low | 84 different post-operative clinical scores (at 3M, 6M, 1Y, 3Y) were used as outcomes, providing a holistic view. | Page 3 / Clinical data |
| Analysis | Low |  | Used a sophisticated pipeline (ANN for compression + SVM for regression) with 50-fold cross-validation or LOOCV | Page 4-5 / Training and validation |
| Overall PROBAST | Low | Low | Extremely robust study due to its large sample size, multimodal approach, and adherence to TRIPOD reporting guidelines. | Final Judgment |
