## Supplementary material for "Machine learning-based neuroimaging for prediction of deep brain stimulation outcomes in movement disorders: Systematic review and meta-analysis": Characteristics of the included studies

Table 1: Characteristics of the included studies.

| Author, Year  Country | Indication for DBS | Number of participants | Age^a^ | Gender^b^ | DBS target | Postoperative outcome | Neuroimaging modality |
| --- | --- | --- | --- | --- | --- | --- | --- |
| Radziunas et al., 2025  Lithuania | PD | 34 | 60 (NP-NP) | 47.1% | STN | Delirium | MRI |
| Chang et al., 2023  China | PD | 38 | 58.9 (7.6) | 44.7% | STN | UPDRS III scores | fMRI |
| Chang et al., 2024  China | PD | 127 | 59.6 (7.4) | 44.9% | STN | UPDRS III improvement | MRI |
| Chen et al., 2021  China | PD | 94 | 63 (NP-NP) | 59.6% | STN | UPDRS III improvement | MRI |
| Chen et al., 2022  China | PD | 98 | 61.9 (7.9) | 55.1% | STN | UPDRS III improvement | fMRI + DTI |
| Diao et al., 2024  China | PD | 138 | 62.1 (8.5) | 52.9% | STN | UPDRS III improvement | MRI |
| Gonzalez-Escamilla et al., 2019  Germany | Dystonia | 51 | 51.3 (13.2) | 51.0% | GPi | Dystonia rating scales improvement^c^ | MRI |
| Haliasos et al., 2024  UK | PD | 120 | 58.7 (7.4) | 67.5% | STN | UPDRS III improvement | MRI |
| Jo et al., 2025  South Korea | PD | 58 | 61 (56-66) | 48.3% | GPi | FOG improvement^d^ | MRI + DTI |
| Li et al., 2025  China | PD | 209 | 64 (NP-NP) | 45.5% | GPi | UPDRS III improvement | MRI |
| Liu et al., 2021  China | PD | 33 | 60 (10.1) | 63.6% | STN | UPDRS III improvement | MRI |
| Peralta et al., 2021  France | PD | 196 | 59.9 (8.1) | 48% | STN/GPi/VIM | H-Y/ UPDRS I-III/ Apathy/ Anxiety/ Depression^e^/ Cognitive function^f^ | MRI |
| Mo et al., 2022  China | PD | 33 | NP | NP | NP | UPDRS III improvement | MRI |
| Ramaraju et al., 2023  India | PD | 34 | NP | NP | STN | UPDRS III improvement | MRI |
| Roberts et al., 2024  USA | PD | 40 | 63.3 (7.5) | 55% | STN | UPDRS III improvement | Quantitative Susceptibility Mapping |
| Saudargiene et al., 2022  Lithuania | PD | 34 | 60.4 (NP-NP) | 47.1% | STN | UPDRS III improvement | MRI |
| Shang et al., 2020  China | PD | 50 | 60.2 (7.8) | NP | STN | UPDRS III improvement | fMRI |
| Wang et al., 2021  USA | PD | 55 | 58.2 (10.1) | NP | STN | UPDRS III improvement | fMRI |
| Yang et al., 2023  China | PD | 44 | 61.2 (10) | 61.4% | STN | UPDRS III improvement | fMRI |
| Younce et al., 2025  USA | PD | 65 | 63.5 (8.6) | 63.1% | STN | UPDRS III improvement | fMRI + MRI |

^a: data are presented as mean (SD) or median (Q1-Q3).
b: Data are presented as male %.
c: Included improvements in Burke–Fahn–Marsden dystonia rating scale and Toronto Western Spasmodic Torticollis Rating Scale.
d: defined as an improvement of >1 points.
e: assessed using the Montgomery-asberg depression rating scale.
f: assessed using a Trail making test, Stroop test, Mattis dementia rating scale, categorical fluency, verbal fluency, and lexical fluency.^
^abbreviations: DBS: Deep Brain Stimulation; PD: Parkinson’s Disease; NP: Not Provided; STN: Subthalamic Nucleus; GPi: Globus Pallidus internus; VIM: Ventral Intermediate Nucleus; FOG: Freezing of Gait;^
